# Deprivation and Early Involuntary Retirement: Area-Level Analysis across English Local Authorities

**DOI:** 10.1101/2021.02.25.21252440

**Authors:** Prosenjit Giri, Subhashis Basu, Abrahjit Giri

## Abstract

**Introduction:** Retirement is a major transition point in life. Falling economic support ratios in many countries have led to a rise in the state retirement age and impending changes to eligibility for state and occupational benefit schemes such as pensions as Governments seek to curb expenditure. Permanently incapacitated individuals from work may face increasing challenges in accessing financial support. Such difficulties may impact those most deprived differentially with potentially significant consequences for morbidity and mortality. Few studies thus far have examined early retirement from a societal perspective. This study explores involuntary retirement in this regard.

**Methods:** A retrospective analysis of the association between deprivation with IHR success rates for Local Authorities (LA) in England between 2015-18 was conducted. Deprivation status was assigned according to the proportion of Lower-Layer Super Output Areas in the most deprived 10% nationally using data from the National Statistics SocioEconomic Classification 2015. Freedom of Information Requests were sent to all 326 LAs in England to obtain data on successful IHR applications, number of active members of respective pensions schemes and numbers of applications.

**Results:** 131 LAs provided complete data for IHR applications, numbers of approved applications and eligible members. Several others provided data on application numbers but not those awarded. he national IHR approval rate was 2.16 per 1000 members, with a range of 0.16 to 8.96. There was a trend towards a greater proportion of approved application per 1000 eligible members in more affluent LAs.

**Conclusion:** The results from this brief analysis suggest that there is an association between increasing rates of ill-health retirement and higher area-level deprivation. Policy should note that those in more deprived areas face a quadruple whammy; a greater risk of becoming incapacitated from public health and occupational exposures, more limited access to medical support, less opportunities for alternative work and potentially disproportionate disadvantage from stringent pension eligibility criteria.

## Main Paper

Retirement is a major transition point in adult life. Those who are in stable employment and remain in good health may have the luxury of choosing their retirement date, but others may be forced into early retirement due to permanent incapacity. There is a considerable body of literature examining the physical, psychological, occupational, and institutional features of involuntary retirement. Objective functional limitations, subjective musculoskeletal complaints, and poor self-rated health are known associations (Pietiläinen et al., 2011; Salonen et al., 2018, Eckhoff et al., 2017). Recurrent back pain predicted early retirement in the Whitehall II Study II of British civil servants in the British Government (Lallukka et al., 2018). Obesity and adverse psychological childhood experiences have also been shown to predict early involuntary retirement (Robroek et al., 2013; Harkonmaki et al., 2019). Amongst Finns, a doubling of the hazard ratio between ill-health and early retirement was identified when a mental health and chronic musculoskeletal disorder were simultaneously present (Kaila-Kangas et al., 2014). Work characteristics including high workload and low job autonomy have been associated with early retirement in several studies, but evidence for other work characteristics such as organisational change, interpersonal conflict, job repetition, peer and managerial support, effort-reward imbalance and organisational injustice appears limited (Björkenstam et al., 2017; Lahelma et al., 2012).

In comparison, there is a relative paucity of literature directly examining demographic and societal factors influencing early involuntary retirement. Financial stability and family security are known to predict early voluntary retirement, but those in well-remunerated high-stakes jobs have favourable conditions to continue working beyond normal retirement age (Visser et al, 2016). A longitudinal analysis of demographic factors in European countries suggests that lower education and single status predict early involuntary retirement (Alavinia et al., 2008). The influence of gender appears to vary, with male manual workers more affected in Scandinavian countries, whereas females appear to be more vulnerable in Korea. Such differences however may reflect occupational demographics rather than an independent risk conferred by gender (Knardhal et al., 2017, Kang et al., 2019).

In many countries, the number of individuals of pensionable age has risen due to a fall in the support ratio of economically active people to retirees. This is compounded by a relative delay in younger individuals entering the labour market. Accordingly, policy changes to encourage older individuals to remain in work have been enacted across Europe, but these have primarily occurred through increases to the national state pension age (Stattin 2005). In the context of work disability, more stringent criteria for eligibility to state and occupational illness benefit schemes have also been introduced. Although many schemes remain generous such as those for public sector workers, these are likely to be reformed further to curb expenditure (Hobson 2019).

Such policy changes are concerning, as the most deprived in the workforce, who are also often the least skilled, may be most affected. This concern not only relates to continuing employability into old age, but also access to financial support when permanently incapacitated from work and the subsequent implications for morbidity and mortality. To our knowledge, few studies have assessed the association between deprivation and early involuntary retirement. In the United Kingdom, data regarding retirees on the grounds of ill-health are captured by several public and private sector institutions. Most private sector schemes cover only a limited number of employees and many in the private sector pertain to specific occupational groups, unlikely to be reflect of a broad demographic. English local authorities (LA) however have broader coverage encompassing a range of occupations, and standardised criteria for early pension eligibility. The aim of this study therefore was to assess the relationship between deprivation and rates of ill-health retirement (IHR) in this setting.

## Methods

In the absence of detailed individual-level data, an analysis was conducted to examine the association between LA area-level deprivation with IHR approvals in England between 2015-18. All 326 LAs in England were ranked according to the proportion of Lower-Layer Super Output Areas in the most deprived 10% nationally using data from the National Statistics Socioeconomic Classification 2015. Rankings are provided for each LA from 1 (least deprived) to 200 (most deprived) [NSSEC 2015]. Accordingly, in this freely available online Microsoft Excel file, 126 LAs have a rank of 200.

The 326 LAs were grouped into quintiles such that those ranked 1-49 were classed as group 1 (most deprived quintile) and those ranked 200 as group 5 (least deprived quintile). A power calculation was conducted to estimate the sample size required to detect a statistically significant difference in rates of approved IHR applications per 1000 active members between the most and least deprived LA quintiles. This was conducted using past data in which a median rate of 4.10 successful applications per 1000 employees (IQR 3.01-6.10) was noted for LAs in England [POOLE 2015]. In the absence of more specific data, we applied these IQR values as estimates of rates of approved IHR in group 1 and 5, respectively. Accepting a probability of a Type 1 error of 0.05, powered at 80%, minimum samples sizes of 7451 would be needed to detect a statistically significant difference between these groups. A Chi-squared test for trend was conducted to examine differences in IHR approvals and applications across deprivation quintiles.

Data on the number of successful applications and active pension scheme members for each year were requested electronically from all 326 LAs under the Freedom of Information Act 2000. E-mail reminders were sent to non-responding LAs after each 28-day period from initial contact, with a maximum of three reminders sent in total. Telephone calls to Freedom of Information Officers at non-responding LAs were also made at the third reminder. Statistical analyses were conducted using Microsoft Excel and SPSS Version 23.0. LAs with no IHR applications during the study period or providing incomplete data were excluded from the analysis. Ethical approval was provided by the Clinical Effectiveness Unit at Sheffield Teaching Hospitals, where the project was registered as a service evaluation.

## Results

131 LAs provided complete data for IHR applications, numbers of successful applications and eligible members. 29 LAs provided partial data, and the remainder either did not respond or requested financial reimbursement which we were unable to offer. The age range for members approved for IHR was from 37 to 65, with a median of 56. Only 4 individuals were retired on the grounds of ill-health under the age of 50, and 3 above the age of 60. The national IHR approval rate was 2.16 per 1000 members, with a range of 0.16 to 8.96.

Table 1 shows the total number of employees eligible to apply for early retirement due to ill-health across the three years with corresponding IHR approval numbers, also presented as a rate per 1000 members. Results are stratified by deprivation quintile. More affluent LAs had higher rates of application success had lower rates of success, with group 4 (2^nd^ least deprived quintile) an outlier. A Chi-squared test was conducted to examine rates of IHR success between the most and least deprived quintiles which was significant at the 95% confidence level (χ^2^ = 780.831, df=1, p<0.05).

**Table 1:** IHR Approvals per 1000 Active Members 2015-18

| Deprivation Quintile | Analysed LAs (N) | Eligible Members (N) | Approved IHR applications (N) | Approved IHR Rate per 1000 members |
| --- | --- | --- | --- | --- |
| 1 | 29 | 448170 | 1220 | 2.72 |
| 2 | 25 | 283063 | 608 | 2.15 |
| 3 | 26 | 259915 | 480 | 1.85 |
| 4 | 22 | 146431 | 268 | 1.83 |
| 5 | 29 | 205329 | 330 | 1.60 |
| Overall | 125 | 1342908 | 2906 | 2.16 |

**Table 2:** IHR Applications per 1000 Active Members 2015-18

| Deprivation Quintile | Analysed LAs (N) | Eligible Members (N) | IHR Applications (N) | IHR Application Rate per 1000 members |
| --- | --- | --- | --- | --- |
| 1 | 16 | 232543 | 904 | 3.89 |
| 2 | 11 | 137500 | 486 | 3.53 |
| 3 | 15 | 108282 | 257 | 2.37 |
| 4 | 11 | 65760 | 151 | 2.29 |
| 5 | 22 | 98554 | 154 | 1.56 |

The study was not powered to detect differences in IHR application rates, and only a modest number of LAs provided raw data for this. Furthermore, of those that did, some were unable to produce corresponding numbers of approved applications and vice-versa. Accordingly, the figures in the Table below, shown for illustration, should be considered a separate dataset from that pertaining to approved applications. Notably however, a trend towards higher application rates in more deprived quintiles was observed.

The collated data also afforded the opportunity to examine the consistency of rates of approved IHRs per 1000 members across individual LAs. The root square mean for approved IHRs was 1.42 per 1000 members, with a range of 0.44 to 2.99. Values for all but 5 LAs lay within two standard errors of the mean, with 2 below and 3 above.

## Discussion

The results from this brief analysis suggest that there is an association between increasing rates of ill-health retirement and higher area-level deprivation. A similar trend was observed for applications, although not formally analysed due to limitations in comparing datasets. The relative consistency of IHR approval rates across English LAs would suggest that variability due to factors such as differing assessment methods and attribution of eligibility criteria, may have a limited role in influencing these results. As discussed, there is a relative paucity of work examining both voluntary and involuntary retirement from a social determinant perspective, but findings here are consistent with those previously (Alavinia, 2008; Radl, 2013). Perhaps the most important message from this work is the expectation that such inequalities may widen with expected future changes in the labour market, and likely further reforms to pension schemes given falling economic support ratios.

Those in more deprived areas face a quadruple whammy; a greater risk of becoming incapacitated from occupational and non-occupational exposures, more limited access to medical support, less opportunities for alternative work and potentially disproportionate disadvantage from stringent pension eligibility criteria. The findings here are illustrative, and if supported by more complete data, could be a broader concern in early involuntary retirement. Where individual-level data regarding demographic variables, deprivation, and related characteristics such as occupation, education level and income are unavailable; a possible crude indicator is ratio of applications to approvals. Unfortunately, the limitations of our dataset precluded meaningful evaluation here, but broadly one may expect this ratio to rise with deprivation status as incapacitated workers may be driven to seek financial support through occupational illness pensions schemes. It is important to remember therefore that policy must address both medical and social determinants of incapacitation due to ill-health, and incapacitation due to job market and labour force changes which render the worker ‘apparently incapacitated’. It is not unreasonable to expect that some employers may use ‘ill-health retirement’ as a means of exiting individuals out of the workforce whom they deem redundant, or unwilling or unable to reskill. Whilst this work provides some early insight into the role of deprivation in influencing early involuntary retirement, there is evidently a need for more detailed analyses, and the first step is to ensure collation of accurate and complete datasets not only at local authority level, but for other state and occupational pensions schemes. This remains a significant limitation of this work, and indeed the findings cannot be used to make inferences about individual risks due to the ecological fallacy. Furthermore, over half of contacted LAs did not respond to our requests.

## Data Availability

Data is publically available at National Statistics SocioEconomic Classification 2015. Department for Communities and Local Government: English Indices of Deprivation 2015.

https://www.gov.uk/government/statistics/english-indices-of-deprivation-2015).

